# Prediction of recurrence and functional status in young ischemic stroke patients: Comparison of machine learning and traditional statistical methods

**DOI:** 10.1101/2025.09.09.25335420

**Authors:** Vinicius Viana Abreu Montanaro, Mina Jacob, Kay Sin Tan, Eleonora Maria de Jesus de Oliveira, Thiago Falcão Hora, Merel S Ekker, Youssra Allach, Mengfei Cai, Karoliina Aarnio, Antonio Arauz, Marcel Arnold, Hee-Joon Bae, Lucrecia Bandeo, Miguel Barboza, Manuel Bolognese, Pablo Bonardo, Raf Brouns, Batnairamdal Chuluun, Enkhzaya Chuluunbatar, Charlotte Cordonnier, Byambasuren Dagvajantsan, Stephanie Debette, Adi Don, Christian Ezinger, Esme Ekizoglu, Simon Fandler-Höfler, Annete Fromm, Thomas Gattringer, Christina Jern, Katarina Jood, Young Seo Kim, Steve J. Kittner, Timothy Kleinig, Catharina JM Klijn, Janika Kõrv, Vinod Kumar, Keon-Joon Lee, Tsong-Hai Lee, Noortje Maaijwee, Nicolas Martinez-Majandar, João P Marto, Man Mohan Mehndiratta, Victoria Mifsud, Gisele Pacio, Vinod Patel, Matthew Phillips, Bartlomiej Piechowski-Jozwiak, Aleksandra Pikula, Jose Ruiz-Sandoval, Bettina von Sarnowski, Richard Swartz, David Tanne, Turgut Tatlisumak, Vincent Thijs, Miguel Viana-Baptista, Rijna Vibo, Teddy Wu, Nilüfer Yesilot, Ullrike Waje-Andreassen, Alessando Pezzini, Anil Tuladhar, Jukka Putaala, Gabriel R de Freitas, Frank-Erik de Leeuw, Rui Li

## Abstract

**Introduction:** Ischemic stroke in young adults is a significant social and economic burden. Machine learning (ML) techniques can potentially predict the outcomes of recurrence and functional status after a stroke more accurately than traditional statistical methods. We sought to predict these outcomes in young individuals with stroke with machine learning and compare that with traditional statistical methods.

**Methods:** This study is part of Global Outcome Assessment Lifelong After Stroke in Young Adults (GOAL) initiative, which collects individual patient data from hospital-based young stroke (18-50 years) cohorts from 29 countries covering all continents worldwide. We compared several common machine learning models with traditional logistic regression to investigate the best models for predicting functional outcome, as measured by the modified Rankin scale at three months post-stroke, and stroke recurrence during follow-up.

**Results:** Functional outcome was available for 7937 patients, and stroke recurrence for 9366 patients. Poor functional outcomes post-stroke occurred in 27.0% of cases, and stroke recurrence in 10.1% of cases during a median follow-up time of 75 months. For functional outcome, multilayer perceptron model achieved the highest mean area under the receiver operating characteristic curve (AUC) at 0.92±0.08. Random forest model attained the highest AUC (0.68±0.03) for predicting stroke recurrence. However, their results were not statistically significantly higher than those for logistic regression.

**Conclusion:** Our work explored the use machine learning to predict outcomes in young stroke patients. However, in our cohort, ML methods provided only moderate added value compared to logistic regression for predicting stroke recurrence and functional outcome.

## Introduction

About 2 million individuals in the 18-50 age group suffer from an ischemic stroke annually worldwide.^1^ Ischemic stroke causes significant social and economic burden, particularly in young patients with a life expectancy of decades ahead. Therefore, prognostic information regarding the recurrence of vascular events and functional outcomes is important.^2^ However, robust studies in large cohorts of patients with a stroke at young age are sparse.^3^ This is partly because these younger patients are usually excluded or underrepresented in larger trials and prognostic studies. In addition, causes of stroke, which are usually a good predictor of recurrence, such as large artery disease and small vessel disease, have a low prevalence in young adults and most likely do not constitute the same prognostic importance for stroke at a younger age, particularly in western countries within a 3-month interval. ^1,4,5,6^

An accurate prediction of the risks of recurrent events and adverse outcomes after a stroke in young adults is potentially key to optimizing personalized treatment and preventing recurrent events. Conventional statistical models such as logistic regression model have been used in clinical research for such kind of prediction tasks ^7–10^. However, compared with traditional statistical models, machine learning (ML) models may capture more complex associations between various risk factors and outcomes,^9–13^ due to their more flexible forms and fewer restrictive assumptions.^11–16^ Consequently, our study sought to explore whether commonly used ML methods could enhance the prediction of recurrent vascular events and functional outcomes in young ischemic stroke patients compared to traditional statistical approaches. The adoption of these ML techniques has the potential to significantly improve our predictive capabilities in clinical practice.

## Methods

### Study Design and Population

The GOAL-initiative is an international multicenter initiative that collects individual patient data from hospital-based young stroke cohorts worldwide, aiming to investigate the risk factors, etiology, and outcome of stroke in young adults. A detailed description of the GOAL-study protocol has been published previously.^2^

Briefly, prospective and retrospective hospital-based cohorts were considered eligible for enrolment if their patients would meet our selection criteria. Cohorts were eligible for inclusion if they recruited consecutive patients aged 18-50 years with a first-ever ischemic stroke. Exclusion criteria included stroke due to traumatic cerebral injury, intracerebral malignancy, cerebral venous thrombosis, iatrogenic stroke due to any medical intervention, or retinal infarct.^2^ Ischemic stroke was defined according to the definition of the World Health Organization (WHO).^3^ We analyzed data from cohorts who contributed data to the GOAL-initiative until June 1^st^, 2020.

### Standard Protocol Approvals, Registrations, and Patient Consents

Before participating, each individual center had to obtain written ethical approval from a local ethical committee for international data sharing. Participating centers were requested to anonymize their data before sending it using a standardized and encrypted electronic sheet containing pre-specified variables of interest, according to the current laws and legislation concerning research conduct at each participating center, to the GOAL-research team of the Neurology department of the Radboudumc, Nijmegen. The key linking anonymized data to individual patients remained at the participating centers. This study was conducted according to the principles of the Declaration of Helsinki (version 60, 19 October 2013) and the Dutch law for human research (WMO). Written informed consent was obtained from all participants in the prospectively collected data, while no informed consent was needed from patients registered in the included retrospective cohorts. Ethical approval was obtained from the Medical Review Ethics Committee region Arnhem-Nijmegen. All data is processed, stored, and will be destroyed after the end of the study according to European Union General Data Protection Regulation.

### Study Data

Study data included demographic characteristics, medical history including medication used on admission, and information on causes and risk factors of ischemic stroke based on diagnostic work-up within 1 month after stroke.

#### Demographics

Date of admission, date of index stroke, age, sex, geographical residence, and self-identified ethnic and racial subgroups including Whites (i.e. non-Hispanic White individuals), Blacks (i.e. non-Hispanic Black individuals), Hispanics, Asians, multi-racial, others or unknown, were registered.

We included patients with ischemic stroke from 29 countries covering all continents except Antarctica. An overview of participating cohorts is available in Supplementary Material Table 1, and the characteristics of all subjects included are summarized in Supplementary Material Table 2. Countries were classified based on their income level at the start of their particular study according to the World Bank Classification in LMICs or HICs (http://datatopics.worldbank.org/world-development-indicators/the-world-by-income-and-region.html).

#### Predictors and Outcome

All variables used for predicting each outcome are listed in Table 1 and 2 respectively. Predictors included vascular risk factors, as stated by the 2014 guidelines of the American Stroke Association^5^, including hypertension, diabetes, dyslipidemia, ever smoking, atrial fibrillation (AF), patent foramen ovale (PFO), and obesity. We also included other risk factors collected (but not required for participation in the GOAL-initiative): migraine, excessive use of alcohol, and illicit drug use. Detailed definitions of collected risk factors are described in the study protocol^2^ and some are summarized in Supplementary Material Table 3 Baseline medications were defined by the premorbid use of these drugs. The predictor, number of risk factors, considered classical vascular risk factors: hypertension, diabetes, dyslipidemia, and smoking^2^. Cohorts that applied a different definition for a given risk factor that could not be reclassified according to GOAL definitions were excluded from that particular sub-analysis. The cause of stroke was classified according to the Trial of Org 10172 In Acute Stroke Treatment (TOAST) criteria.^6^ As part of the preprocessing stage, nominal variables were one-hot-encoded^17^ and continuous variables were standardized.

**Table 1:** Variables used for predicting functional outcome at 3 months post stroke.

| **Modified Rankin Scale at 3 months** | **≤2(n=6837)** | **>2(n=2529)** | **p-value** |
| --- | --- | --- | --- |
| Age in years | 41.0 (7.4) | 41.6 (7.8) | 0.657 |
| Income classification |  |  | **<0.001** |
| HIC | 5832(85.3%) | 1517(60.0 %) |  |
| LMIC | 1005(14.7%) | 1012(40.0 %) |  |
| Ethnicity |  |  | **<0.001** |
| White | 532(7.8%) | 329(13%) |  |
| Black | 26(0.4%) | 7(0.3%) |  |
| Hispanic | 737(10.8%) | 732(28.9%) |  |
| Asian | 4743(69.4%) | 966(38.2%) |  |
| Other | 0(0.0%) | 7(0.3%) |  |
| Unknown | 799(11.7%) | 488(19.3%) |  |
| Season of event |  |  | **0.038** |
| Spring | 2134(31.2%) | 706(27.9%) |  |
| Summer | 1634(23.9%) | 601(23.8%) |  |
| Autumn | 1654(24.2%) | 601(23.8%) |  |
| Winter | 1415(20.7%) | 620(24.5%) |  |
| Gender |  |  | **0.006** |
| Female | 2352(34.4%) | 948(37.5%) |  |
| Infection | 355(5.2%) | 278(11.0%) | **<0.001** |
| Diabetes | 1087(15.9%) | 460(18.2%) | **0.008** |
| Hypertension | 2673(39.1%) | 928(36.7%) | **0.034** |
| Dyslipidemia | 2037(29.8%) | 753(29.8%) | 0.952 |
| Peripheral vascular disease | 47(0.7%) | 53(2.1%) | **<0.001** |
| Ever smoking | 3664(53.6%) | 1294(51.2%) | **0.042** |
| Family history of stroke | 1353(19.8%) | 323(12.8%) | **<0.001** |
| Obesity | 587(8.6%) | 230(9.1%) | 0.444 |
| Migraine | 1880(27.5%) | 237(9.4%) | **<0.001** |
| Hormonal replacement | 519(7.6%) | 141(5.6%) | **0.019** |
| Illicit recent drug use | 211(3.1%) | 53(2.1%) | 0.133 |
| Excessive alcohol use | 929(13.6%) | 452(17.9%) | **0.001** |
| Coronary artery disease | 205(3.0%) | 141(5.6%) | **<0.001** |
| Patent foramen ovale | 690(10.1%) | 194(7.7%) | **0.005** |
| Atrial Fibrillation | 253(3.7%) | 169(6.7%) | **<0.001** |
| Statin baseline | 608(8.9%) | 316(12.5%) | **<0.001** |
| Anticoagulants baseline | 82(1.2%) | 136(5.4%) | **<0.001** |
| Anti platelets baseline | 731(10.7%) | 440(17.4%) | **<0.001** |
| Anti hypertensives baseline | 1374(20.1%) | 662(26.2%)) | **<0.001** |
| Stroke Etiology (TOAST criteria) |  |  | **<0.001** |
| Large vessel disease | 1367(20.0%) | 207(8.2%) |  |
| Small vessel disease | 1593(23.3%) | 579(22.9%) |  |
| Cardioembolic | 957(14.0%) | 478(18.9%) |  |
| Other determined | 1244(18.2%) | 488(19.3%) |  |
| Undetermined | 1676(24.5%) | 776(30.7%) |  |
| Baseline NIHSS (mean) | 3.92(4.58) | 9.78(7.79) | **<0.001** |
| Discharge mRS score |  |  | **0.008** |
| 0 | 1805(26.4%) | 74(2.9%) |  |
| 1 | 2393(35.0%) | 182(7.1%) |  |
| 2 | 1689(24.7%) | 254(10.1%) |  |
| 3 | 649(9.5%) | 6310(28.1%) |  |
| 4 | 247(3.6%) | 774(30.6%) |  |
| 5 | 54(0.8%) | 535(21.2%) |  |
| Number of risk factors |  |  | 0.173 |
| 0 | 1587(23.2%) | 647(25.6%) |  |
| 1 | 2373(34.7%) | 838(33.1%) |  |
| 2 | 1778(26.0%) | 632(25.0%) |  |
| 3 | 847(12.4%) | 332(13.1%) |  |
| 4 | 252(3.7%) | 80(3.2%) |  |

HIC: High income countries; LMIC: Low middle income countries, mRs: Modified Rankin scale; NIHSS: National Institute of Health Stroke Scale; TOAST: trial of org 10172 in acute stroke treatment; NOAC: New oral anticoagulants. Continuous variables are presented as mean (standard deviation), while categorical variables are displayed as n (%), with n representing the count or frequency of the positive class for binary variables.

**Table 2:** Variables used for predicting recurrent stroke during follow-up.

| Stroke recurrence | No(n=7134) | Yes(n=803) | p-value |
| --- | --- | --- | --- |
| Age in years | 40.7(7.7) | 40.3 (7.7) | 0.795 |
| Follow up duration in months | 60.22(68.2) | 92.40 (76.8) | **0.023** |
| Ethnicity |  |  | 0.116 |
| White | 3908(54.8%) | 460(57.3%) |  |
| Black | 262(3.7%) | 36(4.5%) |  |
| Hispanic | 827(11.6%) | 78(9.7%) |  |
| Asian | 641(9.0%) | 78(9.7%) |  |
| Other | 28(0.4%) | 0(0.0%) |  |
| Unknown | 1468(20.6%) | 151(18.8%) |  |
| Income classification |  |  | **0.049** |
| HIC | 5800(81.3%) | 676(84.2%) |  |
| LIMC | 1334(18.7%) | 127(15.8%) |  |
| Season of event |  |  | 0.706 |
| Spring | 2227(31.2%) | 224(27.9%) |  |
| Summer | 1707(23.9%) | 191(23.8%) |  |
| Autumn | 1728(24.2%) | 191(23.8%) |  |
| Winter | 1472(20.6%) | 197(24.5%) |  |
| Gender |  |  | 0.210 |
| Female | 3046(42.7%) | 323(40.3%) |  |
| Infection | 521(7.3%) | 62(7.8%) | 0.763 |
| Diabetes | 692(9.7%) | 88(11.0%) | 0.268 |
| Hypertension | 2340(32.8%) | 275(34.3%) | 0.403 |
| Dyslipidemia | 2554(35.8%) | 312(38.9%) | 0.084 |
| Peripheral vascular disease | 85(1.2%) | 17(2.1%) | 0.104 |
| Ever smoking | 3310(46.4%) | 359(44.7%) | 0.374 |
| Family history of stroke | 1405(19.7%) | 159(19.9%) | 0.914 |
| Obesity | 1227(17.2%) | 135(16.8%) | 0.808 |
| Migraine | 1562(21.9%) | 136(17.0%) | **0.025** |
| Hormonal replacement | 863(12.1%) | 70(8.8%) | **0.028** |
| Illicit recent drug use | 321(4.5%) | 27(3.4%) | 0.310 |
| Excessive alcohol use | 777(10.9%) | 106(13.2%) | 0.071 |
| Coronary artery disease | 321(4.5%) | 39(4.9%) | 0.633 |
| Patent foramen ovale | 1569(22.0%) | 154(19.2%) | 0.160 |
| Atrial Fibrillation | 264(3.7%) | 25(3.2%) | 0.441 |
| Statin baseline | 1234(17.3%) | 139(17.4%) | 0.930 |
| Anticoagulants baseline | 549(7.7%) | 122(15.2%) | **<0.001** |
| Anti platelets baseline | 1206(16.9%) | 174(21.4%) | **0.013** |
| Anti hypertensives baseline | 1305(18.3%) | 170(21.2%) | 0.152 |
| Stroke Etiology (TOAST criteria) |  |  | 0.653 |
| Large vessel disease | 837(11.7%) | 87(10.8%) |  |
| Small vessel disease | 779(10.9%) | 104(13.0%) |  |
| Cardioembolic | 1714(24.0%) | 169(21.0%) |  |
| Other determined | 2127(29.8%) | 253(31.5%) |  |
| Undetermined | 1677(23.5%) | 190(23.7%) |  |
| Baseline NIHSS | 5.75(6.3) | 5.71(6.0) | 0.880 |
| Discharge mRS score |  |  | 0.568 |
| 0 | 1113(15.6%) | 125(15.5%) |  |
| 1 | 1641(23.0%) | 176(21.9%) |  |
| 2 | 1719(24.1%) | 176(21.9%) |  |
| 3 | 1199(16.8%) | 166(20.6%) |  |
| 4 | 884(12.4%) | 87(10.9%) |  |
| 5 | 556(7.8%) | 68(2.4%) |  |
| Number of risk factors |  |  | 0.742 |
| 0 | 1982(27.8%) | 238(29.6%) |  |
| 1 | 2503(35.1%) | 258(32.1%) |  |
| 2 | 1725(24.2%) | 178(22.2%) |  |
| 3 | 762(10.7%) | 106(13.2%) |  |
| 4 | 162(2.3%) | 23(2.9%) |  |

HIC: High income countries; LMIC: Low middle income countries, mRs: Modified Rankin scale; NIHSS: National Institute of Health Stroke Scale; TOAST: trial of org 10172 in acute stroke treatment; NOAC: New oral anticoagulants. Continuous variables are presented as mean (standard deviation), while categorical variables are displayed as n (%), with n representing the count or frequency of the positive class for binary variables.

The prediction outcomes were: 1) poor functional outcome 3 months after index stroke, defined as modified Rankin scale (mRs) > 2; 2) stroke recurrence, defined as subsequent ischemic stroke during follow-up.

### Statistical and ML analysis

For each of the two prediction outcomes, Student’s t-test and Mann-Whitney test were used for comparing the continuous variables between those with or without the outcome. Categorical variables were expressed as proportions and compared between groups with the chi-square test or Fisher’s exact testP values ≤ 0.05 were considered significant.

We compared the logistic regression (LR) model with the following five ML models in predicting both stroke recurrence and functional status 3 months post-stroke:

1. **Decision tree:** this algorithm splits data into binary categories using progressive iterations. It aims to find the optimal features at which to perform data splitting, creating a branching tree-shaped diagram. Each node represents a point at which the data are split, and the leaves at the end of the tree are the output variables.^19^
2. **Support vector machine (SVM):** it classifies training examples into two categories by identifying a hyperplane that best separates the data points. Unlike LR, the optimal hyperplane is positioned to maximize its distance to the closest datapoints from each class, known as the support vectors. The algorithm is tasked with finding the support vectors and hyperplane coefficients. The function can then classify new datapoints falling on either side of the hyperplane.^20^
3. **Multilayer perceptron (MLP):** a type of fully connected neural network modelled after the layer-like histologic stratification of neurons. It consists of an input layer, an output layer, and one or more hidden layers in between performing successive transformations of the input data. During training, the model learns by adjusting the weights of the connections between neurons through backpropagation, progressively refining the transformations to minimize the error between the predicted and actual outputs.
4. **Random forest (RF):** A random forest model is an ensemble model consisting of typically hundreds to thousands of decision trees. It introduces randomness in the construction of decision trees and aggregates the predictions of numerous decision trees to reach a final prediction. It can tackle both classification and regression challenges^18,21^.
5. **Naïve Bayes**: This is a generative ML classifier, which aims to model the distribution of predictors for a specific class. In contrast to discriminative classifiers such as logistic regression, it doesn’t learn the relative importance among predictors for making classification^21^.

We grouped our cohorts by geographical continent into 4 distinct clusters: America, Europe, Asia, and others (Oceania and Africa). Due to the small sample size of the African cohort, we merged it with the Oceanian cohorts, so that we have sufficient samples and outcomes within each group. Using these 4 groups, we conducted internal-external cross-validation (IECV), which is an evaluation framework assessing the model’s generalizability to new data collected independently from the development data, and is particularly suitable for multi-cohort studies like GOAL^22^. In this process, each cluster took turns to be the test set, while the remaining three clusters were used for model training and hyperparameter optimization with auto-WEKA^23^. Models were compared on their test performance.

As stroke recurrence was a rare event (only 10% had recurrence overall) leading to a severe imbalance between the two classes with and without stroke recurrence, we randomly downsampled the majority class to achieve a 1:1 ratio between the majority and minority classes in the training stage within each cross-validation split for all models. This was to ensure that the minority class was not overlooked during training, so that the trained models could have a more balanced performance between the two classes.

We evaluated and compared the test accuracy, sensitivity, specificity, negative and positive predictive values (NPV and PPV), F1 scores and area under receiver operating characteristic curve (ROC-AUC) for all models. Paired Student’s t-test was used to assess the significance of the difference in test metrics between each ML model and logistic regression. We followed the TRIPOD guidelines for prediction of prognosis in populational studies.

SPSS version 24 was used for descriptive analysis and correlation tests, and WEKA version 3.8.6 software was used for running the machine learning and logistic regression models and the comparison between them.

### Data Availability

Data from the GOAL substudy including data supporting the findings of this study will be available from the corresponding author upon reasonable request, after the consent of all GOAL participating centers and approval of local institutional review boards.

## Results

### Cohort Characteristic**s**

Of the total of 17263 patients included in the GOAL-study, for 9366 subjects the functional status measured by the mRs at 3 months after stroke was available, and 2529/9366 (27.0%) had a mRs score >2 after 3 months. For 7937 patients, information regarding stroke recurrence was available, and 803/7937 (10.1%) patients had recurrent stroke during a median follow-up of 75 months. Mean age was 41.0 years (SD 4) and 55% were male; men were older than women (41.5 [SD 7.2] vs 39.7 [SD 7.9] years; p < 0.001) overall. 38.2% were Asian, 30.0% white, 10.2% were Hispanic and 2.5% black. The difference between those who had poor or good functional outcome 3 months post stroke in the entire sample is depicted in Table 1, and difference between those who experienced stroke recurrence and those who did not in Table 2. Similar results of the characteristics of the subjects in each IECV cluster are reported in Supplementary Materials tables 4 and 5.

### Results for Functional Outcome Prediction

Table 3 depicts the average test performance of all models when predicting functional outcome. MLP achieved the highest mean AUC of 0.92±0.08, which was followed closely by RF (0.91±0.09). RF, on the other hand, achieved much higher F1 score (RF=0.85±0.08 vs MLP=0.75±0.10) and PPV (RF=0.76±0.05 vs MLP=0.68±0.10) than MLP. Comparing each ML model against logistic regression, however, no ML model significantly outperformed logistic regression on AUC. SVM was the only ML model that achieved an AUC significantly worse than logistic regression (SVM 0.74±0.07; p-value=0.045).

**Table 3:** Comparison of test sensitivity, specificity, accuracy, and area under curve between logistic regression and machine learning models in predicting functional outcome at 3 months post stroke.

| Models | Logistic Regression | Decision Trees | Support Vector Machine | Multilayer Perceptron | Random Forest | Naive Bayes |
| --- | --- | --- | --- | --- | --- | --- |
| AUC | 0.90 ±0.10 | 0.75 ±0.07 | 0.74 ±0.07* | 0.92 ±0.08 | 0.91 ±0.09 | 0.88 ±0.10 |
| Sensitivity | 0.79 ±0.05 | 0.60±0.04* | 0.76±0.02* | 0.76 ±0.06 | 0.76±0.03 | 0.64 ±0.05 |
| Specificity | 0.85 ±0.07 | 0.84 ±0.11 | 0.84 ±0.03* | 0.89 ±0.02* | 0.87 ±0.08 | 0.87 ±0.07 |
| Accuracy | 0.85 ±0.08 | 0.76 ±0.08 | 0.84 ±0.10 | 0.84 ±0.03 | 0.87 ±0.07 | 0.81 ±0.04 |
| PPV | 0.65 ±0.06 | 0.54 ±0.09 | 0.52 ±0.04 | 0.68 ±0.10 | 0.76 ±0.05 | 0.64 ±0.08 |
| NPV | 0.92 ±0.08 | 0.89 ±0.03 | 0.92 ±0.05 | 0.92 ±0.04 | 0.92 ±0.07 | 0.87 ±0.09 |
| F1 score | 0.78 ±0.05 | 0.81 ±0.07 | 0.78 ±0.04 | 0.75 ±0.10 | 0.85 ±0.08 | 0.81 ±0.09 |

PPV: positive predictive value, NPV: negative predictive value, AUC: Area under ROC curve
\* denotes where the result for an ML model is significantly ( $p \leq 0.05$ ) different from that for the logistic regression model

### Results for Stroke Recurrence Prediction

Table 4 depicts the average test performances of all models in predicting stroke recurrence. The ML model with the highest mean AUC (0.68±0.03) was random forest. Despite having moderate results overall, RF had superior mean sensitivity (0.67±0.04), accuracy (0.65±0.02), AUC (0.68±0.03), F1 score (0.63±0.01) and negative predictive value (0.67±0.04), compared with logistic regression (sensitivity=0.60±0.05, accuracy=0.64±0.03, AUC=0.66±0.03, F1 score=0.60±0.04 and negative predictive value=0.62 ±0.04), However, the differences did not achieve statistical significance (Table 4). MLP, on the other hand, could barely predict stroke recurrence as reflected by its mean AUC (0.55±0.05).

**Table 4:** Comparison of test sensitivity, specificity, accuracy and area under ROC curve between logistic regression and machine learning models in predicting stroke recurrence.

| Models | Logistic Regression | Decision Trees | Support Vector Machine | Multilayer Perceptron | Random Forest | Naive Bayes |
| --- | --- | --- | --- | --- | --- | --- |
| AUC | 0.66.±0.03 | 0.64 ±0.05 | 0.56 ±0.08 | 0.55 ±0.05* | 0.68 ±0.03 | 0.63 ±0.04 |
| Sensitivity | 0.60 ±0.05 | 0.61 ±0.02* | 0.52 ±0.04* | 0.53 ±0.04* | 0.67 ±0.04 | 0.61 ±0.03* |
| Specificity | 0.67 ±0.02 | 0.59 ±0.03* | 0.53 ±0.05* | 0.57 ±0.03* | 0.62 ±0.05 | 0.59 ±0.05 |
| Accuracy | 0.64 ±0.03 | 0.60 ±0.04 | 0.54 ±0.06* | 0.55 ±0.06 | 0.65 ±0.02 | 0.59 ±0.02* |
| PPV | 0.62 ±0.02 | 0.63 ±0.01 | 0.58 ±0.03* | 0.54 ±0.01 | 0.62 ±0.01 | 0.59 ±0.08 |
| NPV | 0.62 ±0.04 | 0.60 ±0.02 | 0.54 ±0.07 | 0.53 ±0.02 | 0.67 ±0.04 | 0.60 ±0.07 |
| F1 score | 0.60 ±0.04 | 0.59 ±0.05 | 0.60 ±0.02 | 0.62 ±0.03 | 0.63 ±0.01 | 0.59 ±0.01 |

PPV: positive predictive value, NPV: negative predictive value, AUC: Area under ROC curve, \* denotes where the result for an ML model is significantly ( $p \leq 0.05$ ) different from that for the logistic regression model

## Discussion

Our study compared several common machine learning methods with logistic regression in predicting functional outcomes and recurrence risk in young stroke patients. MLP model achieved the highest mean AUC in predicting functional outcome, whereas RF attained the highest mean AUC in predicting stroke recurrence. However, the differences between their results and that of logistic regression was not statistically significant for either analysis. Moreover, the highest mean test AUC for predicting stroke recurrence was only 0.68, illustrating the difficulty of this prediction task.

Logistic regression is a linear model that is easy to interpret, and the coefficients can be used to determine the relative importance of different predictors in the model. In contrast, for many ML algorithms such as random forest, it is more difficult to interpret how the predictors interact with each other and the relationship between the predictors and the outcome^24^. Therefore, given also the similarity in AUC, sensitivity, and specificity between the ML methods and logistic regression, logistic regression could be favored in clinical practice in predicting the outcomes.

Our work has similar results to other papers, such as one predicting dementia in cerebral small vessel disease, in which there were no significant differences between ML and traditional statistical models in predictive power^25^. Also, another paper concerning prognostication in intracerebral gliomas found similar results^20^. A systematic review also demonstrated no superiority of ML over logistic regression for clinical prediction models.^26^ Some other clinical studies with similar designs, however, did report superior performance of ML models than statistical models. For example, in a recent study predicting mortality and prolonged hospital stay of stroke patients in the intensive care unit, the Random Forest algorithm was reported to outperform all other ML and conventional models^24^. However, the differences in test AUC between RF and LR were only 0.02-0.03, and the significance of these differences were not assessed. Moreover, although their prediction outcomes were also rare events (e.g., 8% mortality), they did not address the class imbalance problem (having much fewer subjects experiencing the event than those who did not) as we did in the stroke recurrence prediction analysis, which resulted in their models having low sensitivity and could also contribute to the differences between our results.

The strengths of our work include the use of a multi-country cohort covering all continents, which is the largest cohort to date analyzing stroke recurrence and functional outcomes in young stroke patients. We also performed internal-external cross validation, which ensured that the same cohort did not appear in both the internal training and external testing sets, allowing us to test the model’s generalizability to a new population from countries distinct from those in the internal dataset^27,28^.

A limitation of our study is the retrospective nature of data collection. However, our work is currently the attempt with the largest and most diverse young stroke population to generate a prediction model for recurrence and functional outcome in young patients with stroke^8–16^ Additionally, features such as visceral obesity, serum glucose levels, high temperature and psychosocial factors may be related to stroke recurrence and functional outcomes as well,^29,30^ but were not included in this study due to unavailability. The lack of high-dimensional data (e.g., imaging, omics) also prevented us from using more advanced deep learning models. In conclusion, our work highlights the potential use of ML in predicting functional outcomes and recurrence in young stroke patients. However, ML models brought very limited improvement over conventional logistic regression in our experiments. Also given the difficulties in the implementation and interpretation of ML in clinics at this stage, logistic regression may be a preferred method for predicting stroke recurrence and functional outcome due to its simplicity, interpretability, and efficiency.

## Data Availability

All data produced in the present study are available upon reasonable request to the authors

## Declarations

The authors did not receive support from any organization for the submitted work. The authors have no competing interests to declare that are relevant to the content of this article.

## References

1. Ekker MS, Jacob MA, van Dongen MM, et al. Global Outcome Assessment Life-long after stroke in young adults initiative-the GOAL initiative: study protocol and rationale of a multicentre retrospective individual patient data meta-analysis. BMJ Open 2019; 9(11): e031144.

2. Putaala J, Metso AJ, Metso TM, et al. Analysis of 1008 consecutive patients aged 15 to 49 with first-ever ischemic stroke: the Helsinki young stroke registry. Stroke 2009; 40(4): 1195–203.

3. Aho K, Harmsen P, Hatano S, Marquardsen J, Smirnov VE, Strasser T. Cerebrovascular disease in the community: results of a WHO collaborative study. Bull World Health Organ 1980; 58(1): 113–30.

4. Ekker MS, Boot EM, Singhal AB, et al. Epidemiology, aetiology, and management of ischaemic stroke in young adults. Lancet Neurol 2018; 17(9): 790–801.

5. Tan KS, Navarro JC, Wong KS, et al. Clinical profile, risk factors and aetiology of young ischaemic stroke patients in Asia: A prospective, multicentre, observational, hospital-based study in eight cities. Neurology Asia 2014; 19(2) : 117 – 127

6. Adams HP, Jr., Bendixen BH, Kappelle LJ, et al. Classification of subtype of acute ischemic stroke. Definitions for use in a multicenter clinical trial. TOAST. Trial of Org 10172 in Acute Stroke Treatment. Stroke 1993; 24(1): 35–41.

7. Vodencarevic A, Weingartner M, Caro JJ, et al. Prediction of Recurrent Ischemic Stroke Using Registry Data and Machine Learning Methods: The Erlangen Stroke Registry. Stroke. 2022;53:2299–2306

8. Dilsizian ME, Siegel EL. Machine meets biology: a primer on artificial intelligence in cardiology and cardiac imaging. Curr Cardiol Rep. 2018;20(12):139.

9. Holodinsky JK, Yu AYX, Kapral MK, Austin PC. Comparing regression modeling strategies for predicting hometime. BMC Med Res Methodol 2021; 21:138

10. Jones DT, Kerber KA. Artificial Intelligence and the Practice of Neurology in 2035. Neurology. 2022;98:238–245

11. Johnson KW, Soto JT, Glicksberg BS, Shameer K, Miotto R, Ali M, et al. Artificial Intelligence in Cardiology. J Am Coll Cardiol. 2018;71(23):2668–79.

12. Krittanawong C, Zhang H, Wang Z, Aydar M, Kitai T. Artificial intelligence in precision cardiovascular medicine. J Am Coll Cardiol. 2017;69(21):2657–64.

13 Massalha S, Clarkin O, Thornhill R, Wells G, Chow BJW. Decision support tools, systems, and artificial intelligence in cardiac imaging. Can J Cardiol. 2018;34(7):827–38.

14. Mesquita CT. Artificial intelligence and machine learning in cardiology – a change of paradigm. Int. J. Cardiovasc. Sci. 2017;30(3):187–8

15 Moore J. The Dartmouth College Artificial Intelligence Conference: The Next Fifty Years. AI Magazine. 2006;27(4):87–91.

16 De Souza Filho EM, Fernandes FA, De A, Soares CL, et al. Inteligência artificial em cardiologia: conceitos, ferramentas e desafios “quem corre e o cavalo, você precisa ser o joquei -” inteligência artificial em cardiologia. artificial intelligence in cardiology. Arq Bras Cardiol 2019;114 (4):719–725

17 Hancock J, Khoshgoftaar T. Survey on categorical data for neural networks. J Big Data 2020;7:28

18 Chavva RI, Crrawford AL, Mazurek MH, Yuen MM, prabhat AM, Payabvash S et al. Deep Learning Applications for acute Stroke Management. Ann Neurol 2022;00:1–14

19 Holodinski JK, Yu AYX, Kapral MK and Austin PC. Comparing regression modeling strategies for predicting hometime. BMC Med Res Methodol (2021) 21:138

20 Panesar S, D’souza Rhett, Yeh FC et al. Machine Learning Versus Logistic Regression Methods for 2-Year Mortality Prognostication in a Small, Heterogeneous Glioma Database. World Neurosur. Volume 2, April 2019, 100012

21 James G, Witten D, Hastie T, Tibshirani R. An Introduction to Statistical Learning. Springer, 2013 ISSN 1431-875X

22 Takada T, Nijman S, Denaxas S, et al. Internal-external cross-validation helped to evaluate the generalizability of prediction models in large clustered datasets. Journ Clin Epidem. 2021(137):83–91

23 Kotthoff L, Thornton C, Hoos HH, et al. Auto-WEKA2.0:Automaticmodelselection andhyperparameteroptimizationinWEKA. Journ of Mach Learn Reas. 2016 (17):1–5

24 Kurtz P, Peres IT, Soares M, Salluh JIF, Bozza FA. Hospital Length of Stay and 30-Day Mortality Prediction in Stroke: A Machine Learning Analysis of 17,000 ICU Admissions in Brazil. Neurocrit Care. 2022. doi:10.1007/s12028-022-01486-3

25 Li R, Harshfield EL, Bell S, Burkhart M et al. Predicting incident dementia in cerebral small vessel disease: comparison of machine learning and traditional statistical models. Cereb Circ Cogn Behav. 2023 Aug 9;5:100179

26 Christodolou E, Ma J, Collins GS, Steyerberg EW, Verbakel JY, Calster BV. A systematic review shows no performance benefit of machine learning over logistic regression for clinical prediction models. Journ Clin Epidem. 2019 110:12–22

27 Witten IH, Frank E, Hall MA. Data Mining: Practical Machine Learning Tools and Techniques. 3rd ed. San Francisco: Morgan Kaufmann; 2011. p. 191.

28. Collins GS, Dhiman P, Ma, J, et al. Evaluation of clinical prediction models (part 1): from development to external validation. BMJ 2024;384:e074819

29 Balkaya M, Cho S. Optimizing functional outcome endpoints for stroke recovery studies. J Cereb Blood Flow Metab. 2019 Dec; 39(12): 2323–2342

30 Zhang R, Liu H, Pu L. Global Burden of Ischemic Stroke in Young Adults in 204 Countries and Territories. Neurology 2023;100:e422–e434.

