## Supplemental material for "Prediction of recurrence and functional status in young ischemic stroke patients: Comparison of machine learning and traditional statistical methods"

**SUPPLEMENTARY MATERIAL**

Supplemental Table 1: Overview of participating cohorts in alphabetical order

| **Country** | **Population** | **Single centre or Multicentre** | **Number of patients (N)** | **Design** | **Patient selection** | **Stroke definition** | **Study period** |
| --- | --- | --- | --- | --- | --- | --- | --- |
| **Argentina** | Hospital-based | Single centre | 91 | Retrospective | Consecutive | WHO | 1999-2019 |
| **Australia** | Hospital-based | Multicentre | 322 | Retrospective | Consecutive | WHO | 2006-2010 |
| **Austria** | Hospital-based | Single centre | 363 | Retrospective | Consecutive | Radiologically confirmed | 2008-2017 |
| **Belgium** | Hospital-based | Multicentre | 448 | Prospective and Retrospective | Consecutive | WHO | 2007-2008 |
| **Brazil** | Hospital-based | Single centre | 135 | Retrospective | Consecutive | WHO | 2008-2012 |
| **Canada** | Hospital based | Multicentre | 118 | Prospective | Consecutive | WHO | 2013-2016 |
| **Costa Rica** | Hospital-based | Single centre | 166 | Prospective and retrospective | Consecutive | WHO | 2012-2018 |
| **Estonia** | Hospital-based | Multicentre | 424 | Retrospective | Consecutive | WHO | 2003-2012 |
| **Finland** | Hospital-based | Single centre | 1,000 | Retrospective | Consecutive | WHO | 1994-2007 |
| **France** | Hospital-based | Single centre | 307 | Prospective | Consecutive | Radiologically confirmed | 2006-2010 |
| **Germany** | Hospital-based | Single centre | 105 | Prospective and retrospective | Consecutive | Radiologically confirmed | 2007-2012  2017-2019 |
| **India** | Hospital-based | Single centre | 206 | Prospective | Consecutive | WHO | 1988-1997 |
| **Israel** | Hospital-based | Multicentre | 321 | Prospective and retrospective | Consecutive | WHO | 2007-2017 |
| **Italy** | Hospital-based | Multicentre | 2,147 | Prospective | Consecutive | Radiologically confirmed | 2000-2013 |
| **Malaysia** | Hospital-based | Single centre | 179 | Retrospective | Consecutive | Radiologically confirmed | 2016-2017 |
| **Mexico -*Mexico-City*** | Hospital-based | Single centre | 1,456 | Prospective | Consecutive | Radiologically confirmed | 1990-2017 |
| **Mexico - Guadalajara** | Hospital- based | Single centre | 19 | Retrospective and Prospective | Consecutive | Radiologically confirmed | 2017- ongoing |
| **Mongolia** | Hospital-based | Single centre | 148 | Retrospective | Consecutive | WHO | 2012-2017 |
| **The Netherlands** | Hospital-based | Single centre | 451 | Retrospective | Consecutive | WHO | 1980-2010 |
| **New Zealand** | Hospital-based | Single centre | 336 | Retrospective | Consecutive | WHO | 2004-2009 2011-2016 |
| **Norway** | Hospital-based | Single centre | 149 | Prospective | Consecutive | Radiologically confirmed | 2010-2015 |
| **Portugal** | Hospital-based | Single centre | 164 | Prospective and Retrospective | Consecutive | Radiologically confirmed | 2009-2018 |
| **Republic of Korea-**  **Hanyang University** | Hospital-based | Single centre | 160 | Prospective | Consecutive | Radiologically confirmed | 2014-2016 |
| **Republic of Korea-**  **Seoul National University** | Hospital-based | Multicentre  CRCS-K | 5,472 | Prospective | Consecutive | Radiologically confirmed | 2011-2018 |
| **South Africa** | Hospital-based | Single centre | 91 | Retrospective | Consecutive | WHO | 2003-2016 |
| **Sweden** | Hospital-based | Single centre | 552 | Prospective | Consecutive | WHO | 1998-2017 |
| **Switzerland -Bern** | Hospital-based | Multicentre | 428 | Prospective | Consecutive | Radiologically confirmed | 2008-2012 |
| **Switzerland – Luzern** | Hospital- based | Single centre | 50 | Retrospective | Consecutive | Radiologically confirmed | 2016-2018 |
| **Taiwan** | Hospital-based | Single centre | 466 | Retrospective | Consecutive | Radiologically confirmed | 1997-2001 |
| **Turkey** | Hospital-based | Single centre | 100 | Prospective and Retrospective | Consecutive | WHO | 1996-ongoing |
| **United Arab Emirates** | Hospital-based | Single centre | 144 | Prospective | Consecutive | WHO | 2016-2018 |
| **USA** | Hospital-based | Multicentre | 889 | Retrospective | Not Consecutive | WHO | 1992-2008 |

**Supplemental Table 2**: Differences in presence of concurrent risk factors, stratified by demographical characteristics and stroke aetiology

|  | **Total Patients** | **0 Present Risk Factor** | **1 Present Risk Factor** | **2 Present Risk Factors** | **3 Present Risk Factors** | **4 Present Risk Factors** |
| --- | --- | --- | --- | --- | --- | --- |
| **Total reported cases** | **16,845** | **4,419 (26.2)** | **5,530 (34.0)** | **4,148 (24.6)** | **2,034 (12.1)** | **514(3.1)** |
| **Sex**   - **Men** - **Women** | **10,204 (60.6)**  **6,641 (39.4)** | **1,936 (19.0)**  **2,483 (37.4)** | **3,470 (34.0)**  **2,260 (34.0)** | **2,875 (28.2)**  **1,273 (19.2)** | **1,502 (14.7)**  **532 (8.0)** | **421 (4.1)**  **93 (1.4)** |
| **Age-group**   - **18-25** - **26-30** - **31-35** - **36-40** - **41-45** - **46-50** | **894 (5.3)**  **1,070 (6.4)**  **1,721 (10.2)**  **2,804 (16.6)**  **4,748 (28.2)**  **5,608 (33.3)** | **491 (54.9)**  **500 (46.7)**  **658 (38.2)**  **820 (29.2)**  **1,075 (22.6)**  **875 (15.6)** | **333 (37.2)**  **422 (39.4)**  **670 (38.9)**  **988 (35.2)**  **1,627 (34.3)**  **1,690 (30.1)** | **49 (5.5)**  **116 (10.8)**  **278 (16.2)**  **652 (23.3)**  **1,258 (26.5)**  **1,795 (32.0)** | **21 (2.3)**  **28 (2.6)**  **106 (6.2)**  **283 (10.1)**  **625 (13.2)**  **971 (17.3)** | **0 (0)**  **4 (0.4)**  **9 (0.5)**  **61 (2.2)**  **163 (3.4)**  **277 (4.9)** |
| **Ethnicity**   - **Caucasian** - **Black** - **Hispanic** - **Asian** | **5,448 (32.3)**  **499 (3.0)**  **1,700 (10.1)**  **6,566 (39.0)** | **1,559 (28.6)**  **109 (21.8)**  **599 (35.2)**  **1,374 (20.9)** | **1,826 (33.5)**  **156 (31.3)**  **706 (41.5)**  **2,101 (32.0)** | **1,328 (24.4)**  **129 (25.9)**  **256 (15.1)**  **1,831 (27.9)** | **627 (11.5)**  **81 (16.2)**  **112 (6.6)**  **946 (14.4)** | **108 (2.0)**  **24 (4.8)**  **27 (1.6)**  **314 (4.8)** |
| **Geographic location**   - **Oceania** - **Asia** - **Africa** - **Southern Europe** - **Central Europe** - **Northern Europe** - **North America** - **South America** | **578 (3.4)**  **6,510 (38.6)**  **88 (0.5)**  **2,957 (17.6)**  **2,002 (11.9)**  **1,874 (11.1)**  **999 (5.9)**  **1,837 (10.9)** | **126 (21.8)**  **1,347 (20.7)**  **47 (53.4)**  **1,140 (38.6)**  **485 (24.2)**  **375 (20.0)**  **213 (21.3)**  **686 (37.3)** | **188 (32.5)**  **2,087 (32.1)**  **20 (22.7)**  **966 (32.7)**  **748 (37.4)**  **613 (32.7)**  **372 (37.2)**  **736 (40.1)** | **164 (28.4)**  **1,822 (28.0)**  **11 (12.5)**  **552 (18.7)**  **516 (25.8)**  **577 (30.8)**  **236 (23.6)**  **270 (14.7)** | **80 (13.8)**  **946 (14.5)**  **6 (6.8)**  **257 (8.7)**  **218 (10.9)**  **279 (14.9)**  **132 (13.2)**  **116 (6.3)** | **20 (3.5)**  **308 (4.7)**  **4 (4.5)**  **42 (1.4)**  **35 (1.7)**  **30 (1.6)**  **46 (4.6)**  **29 (1.6)** |
| **Country income classes**   - **High income countries** - **Middle income countries** | **13,884 (82.4)**  **2,961 (17.6)** | **3,443 (24.8)**  **976 (33.0)** | **4,648 (33.5)**  **1,082 (36.5)** | **3,579 (25.8)**  **569 (19.2)** | **17,51 (12.6)**  **283 (9.6)** | **463 (3.3)**  **51 (1.7)** |
| **TOAST-Classification**   - **Large vessel atherosclerosis** - **Small vessel disease** - **Cardioembolism** - **Other determined** - **Undetermined** | **2,739 (16.3)**  **2,450 (14.5)**  **3,039 (18.0)**  **3,687 (21.9)**  **4,199 (24.9)** | **366 (13.4)**  **262 (10.7)**  **1,086 (35.7)**  **1,438 (39.0)**  **1,058 (25.2)** | **785 (28.7)**  **671 (27.4)**  **1,089 (35.8)**  **1,412 (38.3)**  **1,507 (35.9)** | **887 (32.4)**  **829 (33.8)**  **589 (19.4)**  **623 (16.9)**  **1,060 (25.2)** | **532 (19.4)**  **528 (21.6)**  **237 (7.8)**  **184 (5.0)**  **474 (11.3)** | **169 (6.2)**  **160 (6.5)**  **38 (1.3)**  **30 (0.8)**  **100 (2.4)** |

**Data are n (%). Prevalence shown for presence of zero, one, two, three, or four vascular risk factors by age groups. Vascular risk factors included hypertension, dyslipidaemia, diabetes and smoking.**

**Supplemental Table 3**: Definitions of selected risk factors

| **Variable or risk factor** | **Definition** |
| --- | --- |
| **Hypertension** | A history of hypertension was defined as its presence either in the patients’ medical history, or when identified during admission for the index event after the acute phase within the first month after stroke. Hypertension was defined as the use of antihypertensive medication and/or systolic blood pressure of 140 mm Hg or greater and/or diastolic blood pressure of 90 mm Hg or greater. |
| **Diabetes mellitus** | A history of diabetes was defined as its presence either in the patients’ medical history, or when identified during admission for the index event. Diabetes was defined as the use of diabetic medication and/or a fasting (defined as no caloric intake for at least 8 hours) plasma glucose >7 mmol/L and/or 2-h PG ≥ 11.1 mmol/L during OGTT and/or HbA1C ≥ 6.5% (48 mmol/mol) and/or symptoms of hyperglycaemia or hyperglycaemic crisis and a random glucose >11.1 mmol/L.^32^ |
| **Dyslipidaemia** | A history of dyslipidaemia was defined as its presence either in the patients’ medical history, or when identified during admission for the index event. Dyslipidaemia was defined as use of statins and/or cholesterol level ≥5.0 mmol/L (193 mg/dL) and/or low-density lipoprotein level ≥3.0 mmol/L (116 mg/dL) and/or high-density lipoprotein level <1.0 mmol/L (39 mg/dL) and/or triglyceride level ≥ 1.7 mmol/L (150mg/dL). |
| **Atrial fibrillation (AF)** | A history of AF (chronic/paroxysmal) was defined as its presence either in the patients’ medical history, or when identified during admission for the index event. AF was defined as diagnosis based on ECG-findings. |
| **Patent foramen ovale (PFO)** | A presence of PFO was defined based on documentation in medical records, or when identified during hospitalization for the index event. PFO was defined as PFO with or without atrial septum aneurysm, as identified on TTE or TEE with or without contrast. |
| **Coronary artery disease** | Coronary artery disease included myocardial infarction and/or angina pectoris. A history of myocardial infarction or angina pectoris was defined as its presence either in the patients’ medical history, or when identified during admission for the index event. |
| **Obesity** | Obesity was defined as a body mass index greater than 30 kg/m^2^, measured during admission for the index event or when reported by the patient. |
| **Migraine** | A history of migraine was defined as its presence either in the patients’ medical history or when identified during hospitalization for the index event. Migraine was defined according to the International Headache Society criteria.^33^ |
| **Ever smoking** | Any current or former smoker. |
| **Excessive alcohol use** | Heavy drinking was defined as the consumption of more than 21 units a week for men and 14 units a week for women, identified at admission for the index event. |
| **Illicit recent drug use** | Within the month prior to stroke. |
| **Infection** | During admission or hospitalization period |

Abbreviations: 2-h PG, 2-hours post glucose; OGTT, oral glucose tolerance test; HbA1c, glycosylated haemoglobin, type A1c; TTE, transthoracic echocardiogram; TEE, transoesophageal echocardiogram.

Supplemental material table 4: Characteristics of the cluster of patients used for internal-external validation functional outcome at 3 months post stroke

| IECV cluster | Europe* | | America* | | Asia* | | Others* | |
| --- | --- | --- | --- | --- | --- | --- | --- | --- |
| N | 3796 | | 1498 | | 3840 | | 403 | |
| Outcome | MRs **≤2** | MRs **>2** | MRs **≤2** | MRs **>2** | MRs **≤2** | MRs **>2** | MRs **≤2** | MRs **>2** |
| N per outcome | 2771(73%) | 1025(27%) | 1048(70%) | 450(30%) | 2726(71%) | 1114(29%) | 278 (65%) | 125(35%) |
| Age (mean) | 43.1 | 42.8 | 41.1 | 41.5 | 39.5 | 40.1 | 40.0 | 40.1 |
| Female sex (%) | 39% | 38% | 38% | 36% | 37% | 39% | 33% | 32% |
| Ethnicity  White  Black  Hispanic  Asian  Other  Unknown | 70%  10%  6%  7%  4%  3% | 70%  11%  8%  5%  3%  3% | 40%  12%  34%  7%  4%  3% | 36%  14%  34%  8%  3%  4% | 1%  2%  1%  82%  12%  2% | 1%  4%  1%  81%  11%  2% | 77%  17%  1%  0%  4%  1% | 72%  20%  2%  0%  5%  1% |
| Season of event  Spring  Summer  Autumn  Winter | 33%  25%  20%  22% | 32%  26%  21%  21% | 33%  24%  21%  22% | 30%  26%  22%  22% | 27%  29%  24%  20% | 25%  30%  23%  22% | 28%  28%  23%  21% | 27%  28%  23%  22% |
| Infection | 5% | 4% | 4% | 6% | 5% | 8% | 6% | 7% |
| Diabetes | 15% | 13% | 14% | 17% | 11% | 14% | 14% | 17% |
| Hypertension | 29% | 27% | 27% | 29% | 26% | 27% | 27% | 29% |
| Dyslipidemia | 25% | 22% | 24% | 27% | 25% | 24% | 25% | 26% |
| Peripheral vascular disease | 0.5% | 0.3% | 0.2% | 0.7% | 0.5% | 0.9% | 0.1% | 0.2% |
| Ever smoking | 51% | 54% | 52% | 55% | 40% | 45% | 49% | 52% |
| Family history of stroke | 14% | 12% | 11% | 12% | 15% | 17% | 14% | 16% |
| Obesity | 9% | 7% | 10% | 13% | 7% | 8% | 5% | 7% |
| Migraine | 20% | 22% | 19% | 20% | 18% | 22% | 20% | 22% |
| Hormonal replacement | 8% | 6% | 7% | 6% | 4% | 5% | 4% | 5% |
| Illicit recent drug use | 5% | 5% | 6% | 7% | 7% | 9% | 8% | 9% |
| Excessive alcohol use | 16% | 15% | 17% | 15% | 15% | 17% | 16% | 18% |
| Coronary artery disease | 2% | 2% | 1% | 2% | 3% | 2% | 3% | 3% |
| Patent foramen ovale | 10% | 10% | 12% | 10% | 13% | 11% | 11% | 12% |
| Atrial Fibrillation | 3% | 2% | 1% | 2% | 2% | 2% | 3% | 4% |
| Statin baseline | 7% | 7% | 4% | 7% | 7% | 7% | 8% | 7% |
| Anticoagulants baseline | 2% | 2% | 1% | 2% | 1% | 2% | 1% | 2% |
| Anti platelets baseline | 8% | 9% | 7% | 9% | 8% | 10% | 7% | 8% |
| Anti hypertensives baseline | 18% | 17% | 15% | 14% | 18% | 14% | 19% | 16% |
| Stroke Etiology (TOAST criteria)  Large vessel disease  Small vessel disease  Cardioembolic  Other determined  Undetermined | 20%  22%  15%  23%  20% | 19%  23%  13%  22%  23% | 22%  22%  13%  23%  20% | 20%  22%  14%  21%  23% | 24%  20%  10%  21%  25% | 22%  20%  11%  20%  27% | 20%  20%  10%  20%  30% | 20%  19%  10%  16%  35% |
| Baseline NIHSS (mean) | 4.04 | 3.99 | 3.54 | 3.89 | 3.84 | 4.19 | 4.14 | 4.25 |
| Discharge mRS score  0  1  2  3  4  5 | 25%  33%  22%  10%  7%  2% | 3%  8%  11%  30%  35%  13% | 24%  35%  20%  11%  8%  2% | 4%  7%  12%  31%  36%  10% | 25%  34%  22%  9%  8%  2% | 6%  8%  10%  30%  35%  11% | 33%  25%  22%  9%  8%  3% | 6%  8%  7%  33%  37%  9% |
| Number of risk factors  0  1  2  3  4 | 25%  33%  23%  10%  9% | 27%  29%  25%  15%  4% | 27%  30%  21%  12%  10% | 22%  25%  22%  18%  13% | 29%  30%  20%  11%  10% | 25%  23%  21%  17%  14% | 33%  28%  21%  11%  7% | 29%  25%  21%  14%  11% |

* Europe cluster comprises the following countries/cohorts:Argentina, Austria,Belgium, Estonia, Finland, France, Germany, Italy, Netherlands, Norway, Portugal, Switzerland -Bern, Switzerland – Luzern and Turkey

* America cluster comprises the following countries/cohorts: Brazil, Canada, Costa Rica, Mexico -Mexico city, Mexio – Guadalajara and United States of America

* Asia cluster comprises the following countries/cohorts: India, Israel, Malaysia, Mongolia, Republic of Korea-Hanyang University, Republic of Korea-Seoul National University, Taiwan and United Arab Emirates

* Others cluster comprises the following countries/cohorts: Australia, New Zealand and South Africa

Supplemental material table 5: Characteristics of the cluster of patients used for internal-external validation for predicting recurrent stroke during follow-up

| IECV cluster | Europe* | | America* | | Asia* | | Others* | |
| --- | --- | --- | --- | --- | --- | --- | --- | --- |
| N | 3175 | | 1270 | | 3254 | | 397 | |
| Outcome | **Recurrence: no** | **Recurrence:yes** | **Recurrence:no** | **Recurrence:yes** | **Recurrence:no** | **Recurrence:yes** | **Recurrence:no** | **Recurrence:yes** |
| N | 2856(90%) | 319(10%) | 1140(91%) | 130(9%) | 2927(90%) | 327(10%) | 354(89%) | 43(11%) |
| Age (mean) | 40.1 | 40.8 | 40.2 | 40.9 | 40.5 | 41.1 | 39.9 | 40.1 |
| Follow up duration in months | 62.2(66.2) | 91.4 (75.8) | 60.2(65.2) | 92.4 (76.8) | 63.2(66.2) | 93.4 (77.8) | 59.2(62.2) | 88.4 (72.8) |
| Female sex (%) | 40% | 41% | 41% | 41% | 40% | 41% | 40% | 42% |
| Ethnicity  White  Black  Hispanic  Asian  Other  Unknown | 0%  9%  6%  6%  5%  3% | 0%  10%  9%  5%  3%  3% | 49%  25%  10%  8%  5%  3% | 47%  26%  13%  7%  4%  3% | 1%  8%  1%  81%  6%  3% | 2%  9%  0%  83%  4%  2% | 79%  15%  2%  0%  2%  1% | 75%  19%  1%  0%  1%  2% |
| Season of event  Spring  Summer  Autumn  Winter | 31%  24%  22%  23% | 30%  25%  22%  22% | 29%  26%  21%  24% | 27%  28%  22%  23% | 30%  24%  22%  24% | 28%  28%  21%  23% | 33%  25%  20%  22% | 31%  26%  22%  21% |
| Infection | 6% | 7% | 5% | 7% | 6% | 8% | 8% | 9% |
| Diabetes | 10% | 11% | 10% | 11% | 9% | 10% | 12% | 13% |
| Hypertension | 27% | 28% | 26% | 28% | 25% | 27% | 26% | 27% |
| Dyslipidemia | 30% | 35% | 31% | 35% | 32% | 33% | 30% | 33% |
| Peripheral vascular disease | 0.8% | 1% | 0.8% | 1% | 0.5% | 0.7% | 0.6% | 0.7% |
| Ever smoking | 45% | 47% | 45% | 47% | 49% | 47% | 52% | 55% |
| Family history of stroke | 18% | 19% | 17% | 19% | 18% | 19% | 19% | 20% |
| Obesity | 14% | 12% | 15% | 13% | 10% | 12% | 9% | 11% |
| Migraine | 21% | 22% | 20% | 22% | 20% | 20% | 19% | 20% |
| Hormonal replacement | 8% | 6% | 7% | 6% | 6% | 5% | 4% | 3% |
| Illicit recent drug use | 5% | 4% | 5% | 5% | 4% | 5% | 6% | 7% |
| Excessive alcohol use | 11% | 14% | 10% | 13% | 11% | 13% | 15% | 14% |
| Coronary artery disease | 3% | 4% | 3% | 4% | 2% | 3% | 2% | 3% |
| Patent foramen ovale | 20% | 22% | 21% | 22% | 20% | 22% | 21% | 20% |
| Atrial Fibrillation | 3% | 2% | 2% | 2% | 1% | 2% | 2% | 2% |
| Statin baseline | 15% | 16% | 16% | 17% | 18% | 19% | 17% | 19% |
| Anticoagulants baseline | 7% | 14% | 8% | 16% | 8% | 17% | 7% | 15% |
| Anti platelets baseline | 15% | 20% | 16% | 22% | 15% | 23% | 13% | 22% |
| Anti hypertensives baseline | 18% | 20% | 19% | 22% | 19% | 23% | 14% | 20% |
| Stroke Etiology (TOAST criteria)  Large vessel disease  Small vessel disease  Cardioembolic  Other determined  Undetermined | 12%  10%  25%  30%  23% | 10%  14%  24%  29%  23% | 11%  11%  24%  31%  23% | 9%  15%  23%  30%  23% | 13%  12%  25%  29%  21% | 11%  15%  24%  28%  22% | 15%  11%  22%  28%  24% | 14%  13%  25%  26%  22% |
| Baseline NIHSS (mean) | 5.64 | 5.70 | 5.74 | 5.70 | 5.61 | 5.63 | 5.61 | 5.63 |
| Discharge mRS score  0  1  2  3  4  5 | 15%  25%  24%  16%  12%  8% | 14%  24%  22%  20%  11%  9% | 14%  26%  25%  15%  13%  7% | 15%  25%  21%  19%  12%  8% | 15%  27%  23%  16%  12%  7% | 13%  26%  22%  20%  11%  8% | 15%  27%  23%  16%  12%  7% | 13%  26%  22%  20%  11%  8% |
| Number of risk factors  0  1  2  3  4 | 25%  33%  23%  10%  9% | 27%  29%  25%  15%  4% | 24%  34%  22%  11%  9% | 26%  30%  24%  16%  4% | 25%  33%  22%  11%  9% | 27%  31%  23%  15%  4% | 27%  33%  20%  11%  9% | 29%  31%  23%  12%  5% |

* Europe cluster comprises the following countries/cohorts:Argentina, Austria,Belgium, Estonia, Finland, France, Germany, Italy, Netherlands, Norway, Portugal, Switzerland -Bern, Switzerland – Luzern and Turkey

* America cluster comprises the following countries/cohorts: Brazil, Canada, Costa Rica, Mexico -Mexico city, Mexio – Guadalajara and United States of America

* Asia cluster comprises the following countries/cohorts: India, Israel, Malaysia, Mongolia, Republic of Korea-Hanyang University, Republic of Korea-Seoul National University, Taiwan and United Arab Emirates

* Others cluster comprises the following countries/cohorts: Australia, New Zealand and South Africa
